# Antidepressant and mental health care utilization in pregnant women with depression and/or anxiety: an interrupted time-series analysis

**DOI:** 10.1101/2021.09.29.21264292

**Authors:** Nhung TH Trinh, Hedvig ME Nordeng, Gretchen Bandoli, Malin Eberhard-Gran, Angela Lupattelli

**Affiliations:** PharmacoEpidemiology and Drug Safety Research Group, Department of Pharmacy, and PharmaTox Strategic Research Initiative, Faculty of Mathematics and Natural Sciences, University of Oslo, Oslo, Norway; Department of Child Health and Development, Norwegian Institute of Public Health, Oslo, Norway; Department of Pediatrics, University of California San Diego, La Jolla, CA, USA; Department of Family Medicine and Public Health, University of California San Diego, La Jolla, CA, USA; Norwegian Research Centre for Women’s Health, Women’s and Children’s Division, Oslo University Hospital, Rikshospitalet, Oslo, Norway; Institute for Clinical Medicine, University of Oslo, Oslo, Norway

**Keywords:** antidepressants, pregnancy, mental health care utilization, maternal depression, maternal anxiety, interrupted time-series analysis, k-means trajectory modelling, perinatal pharmacoepidemiology

## Abstract

**Objectives:** To assess mental health care utilization patterns during and after pregnancy in women with depression and/or anxiety in Norway according to antidepressant fill trajectories in pregnancy.

**Method:** We conducted a registry-linkage cohort study of pregnancies within women having outpatient visit for depression and/or anxiety and antidepressant fills in the six months prior to pregnancy identified from four national registries of Norway (2009-2018). Number of consultations for depression/anxiety per 100 pregnancies as proxy of mental health care utilization were modelled using interrupted time-series analysis with first month into pregnancy and first month after delivery as interruption points. We investigated the time window including six months prior to pregnancy, eight months into pregnancy and one year postpartum. Antidepressant fill trajectories in the corresponding time window were identified using longitudinal k-means trajectory modelling.

**Results:** The cohort included 8,460 pregnancies within 8,062 women with depression/anxiety. We observed reduced mental health care utilization when pregnant women entered the course of pregnancy (negative slopes during pregnancy for all psychiatric specialists and psychologists). The declines were observed for all antidepressant fill trajectories (i.e., discontinuers and continuers) except interrupters (i.e., discontinued then resumed treatment). We found increased mental health care utilization in the postpartum year, notably in interrupters (positive slopes in consultation rates with specialists of outpatient clinics and public-contracted psychiatrists)..

**Conclusions:** Pregnancy was associated with reduced mental health care utilization regardless of whether antidepressant treatment was maintained during pregnancy or not. Increases in mental health care utilization were observed in the postpartum year, especially in interrupters.

**SIGNIFICANT OUTCOMES:**

- Pregnancy is associated with reduced mental health care utilization in pregnant women with depression and/or anxiety in Norway.
- The reduction in mental health care utilization during pregnancy was observed for all antidepressant fill trajectories, except for interrupters.
- Increased in mental health care utilization in the postpartum year was observed, especially among those who discontinued then resumed antidepressant treatment.

**LIMITATIONS:**

- The reasons for antidepressant discontinuation, and whether that was due to symptom remission were not assessed.
- It was not possible to determine whether reduced mental health care utilization was associated with undertreatment during pregnancy.
- It was not possible to measure directly the use of psychosocial interventions and psychotherapy.

## INTRODUCTION

Perinatal mental health disorders affect a significant number of women with debilitating and potentially life-threatening consequences for both parents and their children.^1–3^ Maternal or perinatal depression is an umbrella term encompassing several different depressive conditions, which require different approaches to treatment. Clinical guidelines recommend psychosocial interventions and psychotherapy as first-line treatment for general and maternal depression.^4^ Pharmacotherapy with an antidepressant is often needed in moderate to severe cases or when the patient does not respond to first-line psychotherapy.^5^

One to five percent of women in Europe fill antidepressant prescriptions during pregnancy, mainly selective serotonin reuptake inhibitors (SSRIs), with depression and anxiety being the predominant indications for use.^6–8^ Antidepressant treatment discontinuation is highly prevalent (approximately 50%) in pregnant women, especially before the time of conception.^9–13^ The discontinuation of antidepressant medications in pregnant women with major depression is associated with risk of relapse and postpartum depression for the mothers and several adverse consequences for the developing neonates.^14–16^ A recent study using interrupted time-series analysis (ITSA) visualized a sudden decline in prescription fills for antidepressants (from 1500 to 500 prescription fills per week) starting two to five weeks into pregnancy among prevalent users in Norway.^12^ The cessation of pharmacologic treatment raises concerns about whether or not antidepressant discontinuers receive adequate alternative and complement care (i.e., psychosocial interventions and psychotherapy) as well as closer psychiatric follow-up after discontinuation. A similar approach examining mental health care utilization using ITSA within pregnant women with similar patterns of antidepressant use may provide better understanding about the impact of pregnancy and antidepressant discontinuation on non-pharmacologic care and psychiatric follow-up.

In a population of pregnancies of women having depression/anxiety, we aimed to assess the changing patterns of mental healthcare utilization using ITSA in the overall sample and by antidepressant trajectories. We used consultation rates for depression/anxiety with psychiatry specialists and psychologists as proxy of mental healthcare utilization.

## METHODS

### Data sources

We conducted a nationwide cohort study based on data from 2009-2018 from the Medical Birth Registry of Norway (MBRN) linked to the Norwegian Prescription Database (NorPD), the Norway Control and Payment of Health Reimbursement (KUHR) and the Norwegian Patient Registry (NPR) using unique personal identification numbers.^17,18^

The MBRN is a population-based registry containing information on all births in Norway since 1967. MBRN is based on mandatory notification of all pregnancies lasting more than 12 weeks. In MBRN, the information available for each pregnancy includes maternal identification, demographic information, information on the mother’s health before and during pregnancy, complications during pregnancy and delivery, date of birth and gestational length and other information on the infants.^18,19^

The NorPD is a nationwide registry on all prescribed medications irrespective of reimbursement, dispensed at pharmacies to individual patients treated in primary care since 2004. The medications are classified according to the Anatomical Therapeutic Chemical (ATC) classification system.^17,20^

The KUHR is an administrative database based on electronically submitted reimbursement claims from healthcare professionals to the Norwegian Economics Administration. KUHR contains information on utilization of healthcare from primary and secondary care (e.g., date of consultation, type of practicing, and diagnostic codes). Diagnostic codes follow the WHO’s International Classification of Diseases (ICD-10) and International Classification of Primary Care (ICPC-2) which is more frequently used by general practitioners.

The NPR is an administrative database of records reported by all government-owned hospitals and outpatient clinics, and by all private health clinics that receive governmental reimbursement. NPR contains information on admission to hospitals and specialist health care on an individual level from 2008. Diagnostic codes in NPR follow the ICD-10.

### Study population

The current study includes pregnancies that meet the following criteria: valid maternal ID registered in MBRN; gestational length ≥ 32 weeks (as done in prior study;^21^ this aims to ensure that all pregnancies had comparable chance of being exposed to antidepressant during pregnancy); pregnancy outcome (both livebirths and stillbirths) between 2009 and 2018; at least one outpatient visit for depression (ICD-10: F32 and F33; ICPC-2: P76) and/or anxiety (ICD-10: F40 and F41, ICPC-2: P74) and at least one AD prescription filled at any time in the six months (i.e., 168 days) prior to pregnancy start. Since the indication of AD were not directly available in the databases, the last two criteria maximize the possibility that antidepressants were filled to treat depression/anxiety. The start of pregnancy was estimated from the last menstrual period date via ultrasound, date of delivery and gestational length (all ascertained in the MBRN). Because one woman may present with more than one pregnancy in the cohort leading to non-independent observation and possible overlap between the post-delivery period of one pregnancy with the pre-pregnancy of the following one, we excluded pregnancies among the same women with an interpregnancy interval less than one year (n=60).

### Mental health care utilization

Since 2008, prescribers in Norway must indicate the reimbursement code for each prescription using the ICD-10 or the ICPC-2 coding system.^20^ Among antidepressant prescription fills available in our dataset where reimbursement codes are available, nearly 90% were issued by general practitioners (GPs). In addition, GPs may only prescribe antidepressants and/or provide referral to psychiatric specialists and/or psychologist for psychotherapy after screening for depression/anxiety. Therefore, we assume that consultations with depression/anxiety (ICD-10: F32, F33, F40, F41 and ICPC-2 codes: P74, P76) as reimbursement code (hereafter referred as consultations for depression/anxiety) seen by psychiatric specialists and by psychologists are mainly for psychotherapy and/or non-pharmacological psychiatric follow-up. Similar definition of mental healthcare utilization has been applied in prior research.^23–25^

We explored the patterns of mental health care utilization using number of consultations for depression/anxiety seen by psychiatric specialists and/or psychologists registered in KUHR including psychiatric specialists/psychologists of outpatient clinics, public-contracted psychiatrists, and public-contracted psychologists. Of note, public-contracted specialists are practicing specialists, in this case psychiatrists or psychologists, who receive an operating subsidy from the public sector. Public-contracted specialists assess and treat conditions that do not require hospitalization, similar to outpatient clinics.^22^

Our main outcome measure was the number of consultations for depression/anxiety seen by psychiatric specialists and/or psychologists per 100 pregnancies, aggregated by 28-day month in the time window including six months prior to pregnancy, eight months into pregnancy and twelve months after birth making a total of 26 time points for the analysis.

### Antidepressant fill trajectories

In the attempt to explore whether the changes in mental health care utilization were associated with antidepressant filling patterns, we extracted information on antidepressant prescription fills (ATC code starting with N06A) from NorPD and modelled antidepressant exposure into longitudinal trajectories. In brief, the expected duration of treatment of each antidepressant dispensation was generated using the PRE2DUP method based on package parameters and clinical guidelines.^26^ Antidepressant exposure by week (coded as 1 if any day of the given week was covered by at least one antidepressant dispensation and 0 otherwise) was modelled using longitudinal k-means trajectory modelling.^21^ We investigated the 108-week time window including 24 weeks prior to pregnancy, 32 weeks into pregnancy, and 52 weeks after delivery. Possible trajectories are labelled as follow: discontinuers (i.e., pregnancies within women who discontinued antidepressant in pregnancy and did not resume the treatment), continuers (i.e., who continued the antidepressant treatment throughout the period), and interrupters (i.e., who discontinued antidepressant in pregnancy and then resumed the treatment after childbirth).

### Data analyses

We examined the changes in mental healthcare utilization during pregnancy and within the first postpartum year using ITSA. The monthly consultation rates for depression/anxiety in the 26-month time window were modelled with segmented linear regression with first month into pregnancy and first month after delivery serving as interruption points. These interruption points were chosen because we hypothesized that pregnancy start and delivery preceded changes in mental healthcare utilization.

The ITSA was done separately for psychiatric specialists/psychologists of outpatient clinics, public-contracted psychologists, and public-contracted psychiatrists, in the overall population and by antidepressant prescription fill trajectories.

### Sensitivity analyses

We conducted a set of sensitivity analyses to test the robustness of our results.

First, we limited our study population to one random pregnancy per woman to obtain a fully independent sample.

Second, we extended the look-back window to retrieve depression/anxiety diagnosis to one year in the inclusion criteria (i.e., having outpatient visit for depression/anxiety within one year prior to pregnancy) because depression/anxiety are known to be long-term illnesses.

Third, we explored the changes in the consultation rate for depression/anxiety with the GPs to contrast this consultation pattern with the different antidepressant prescription fill trajectories. The rationale behind this latter analysis is that the majority of antidepressant prescriptions in pregnant women are issued by GPs.

Fourth, we modelled the changes in the consultation rate for any conditions seen by GPs and specialists of outpatient clinics as done in the main analyses to test whether the observed changes are specific for mental health care utilization.

Data management and statistical analyses were performed with Stata/MP 16.0 and R 4.0.5 for Windows.

## RESULTS

### General description

Among 592,189 pregnancies registered in the MBRN during 2009-2018, a total of 8,460 pregnancies within 8,092 women were included in this study (**Figure S1**). In total, 8,314/8,460 (98.3%) resulted in a live birth. The characteristics of the overall population are presented in **Table 1**.

**Table1.** Characteristics of pregnancies included in the study population, Norway, 2009–2018 (8,460 pregnancies)

| Characteristic | Study population |
| --- | --- |
|  | N (%) |
| **Maternal age** |  |
| ≤24 years | 1,673 (19.8) |
| 25-29 years | 2,587(30.6) |
| 30-34 years | 2,521 (29.8) |
| ≥35 years | 1,679 (19.9) |
| **Marital status** |  |
| Married/cohabiting | 7,167 (84.7) |
| Other | 1,293 (15.3) |
| **Parity** |  |
| 0 | 3,896 (46.1) |
| 1 | 4,564 (53.9) |
| **Plurality** |  |
| Singleton | 8,338 (98.6) |
| Multiple | 122 (1.4) |
| **Obstetric index** □ |  |
| 0 | 5,342 (63.1) |
| 1 | 1,997 (23.6) |
| ≥ 2 | 1,121 (13.3) |
| **Previous miscarriage or stillbirth** |  |
| Yes | 2,329 (27.5) |
| No | 6,131 (72.5) |
| **Smoking before pregnancy** |  |
| Missing | 1,256 (14.9) |
| Yes | 5,024 (59.4) |
| No | 2,180 (25.8) |
| **Pregnancy planning** □ □ |  |
| Potentially yes | 3,063 (36.2) |
| Potentially no | 5,397 (63.8) |
| **Severity of depression/anxiety in six months before pregnancy** □ □ □ |  |
| High | 4,696 (55.5) |
| Low | 3,764 (44.5) |
| **Comorbidity in six months before pregnancy** |  |
| Other psychiatric illnesses than depression and/or anxiety | 1,172 (13.9) |
| Eating disorders | 227 (2.7) |
| Bipolar disorders | 224 (2.7) |
| Thyroid disorders | 199 (2.3) |
| **Comedication in six months before pregnancy** |  |
| Opioid analgesic | 1,102 (13.0) |
| Antiepileptics | 474 (5.6) |
| Antipsychotics | 784 (9.3) |
| Benzodiazepines | 2,019 (23.9) |
| Thyroid hormones | 365 (4.3) |
| **Antidepressant in six months before pregnancy** |  |
| SSRI | 5,761 (68.1) |
| SNRI | 2,261 (26.7) |
| Others | 438 (5.2) |

□ Adapted from Bateman et al., using the variables available in MBRN (age, asthma, pre-gestational diabetes, chronic hypertension, kidney disease, previous caesarean section, multiple gestation) and weighting the variables as done by Bateman et al.
□ □ Reporting use of folic acid before pregnancy (extract from MBRN), filling clomiphene (ATC code G03GB02) before pregnancy (extract from NorPD), discontinuing hormonal contraception for systemic use (ATC-code G03A, excluding contraceptive patches, G03AA13, injections, G03AC06, implants, G03AC08, and emergency contraceptives, G03AD) before pregnancy, having preconception encounters in hospital (ICD 10 – Z30: Encounter for contraceptive management).
□ □ □ Pregnancies with at least one of the following proxies in the year prior to pregnancy were classified as high severity: one or more psychiatric inpatient stay or outpatient specialist encounters with a psychiatric diagnosis (diagnoses with ICD-10: F01-F99 except for F32, F33, F40, F41) or deliberate self-harm diagnoses (ICD-10: X71-X83) as registered in NPR; having filled prescriptions for other nervous system drugs (ATC code N except N06 registered in NorPD)

In the time window from six months prior to pregnancy to one year postpartum, a total of 127,473 visits (average: 17.4 visits/pregnancy) for depression/anxiety were registered in KUHR. Of these, 55,465 visits (43.4%) were seen by GPs, 46,234 (26.2%) by psychiatric specialists and/or psychologists of outpatient clinics, 9,836 (7.7%) by public-contracted psychologists, and 9,544 (7.5%) by public-contract psychiatrists.

### Antidepressant fill trajectories

Based on KML modelling, we clustered our population into four trajectories when modelling antidepressants exposure (**Figure S2**): (A) Late discontinuers (discontinued treatment around the end of pregnancy) – 33.8% (B) Early discontinuers (discontinued treatment around the start of pregnancy) – 30.4% (C) Continuers – 20.6% and (D) Interrupters (discontinued treatment around the end of pregnancy then resumed in postpartum period) – 15.2%.

### Mental health care utilization in the time window from six months prior to pregnancy to one year postpartum

Consultation rates for depression/anxiety with interruption points at first month into pregnancy and first month after delivery are visualized in **Figure 1**.

**Figure 1.**
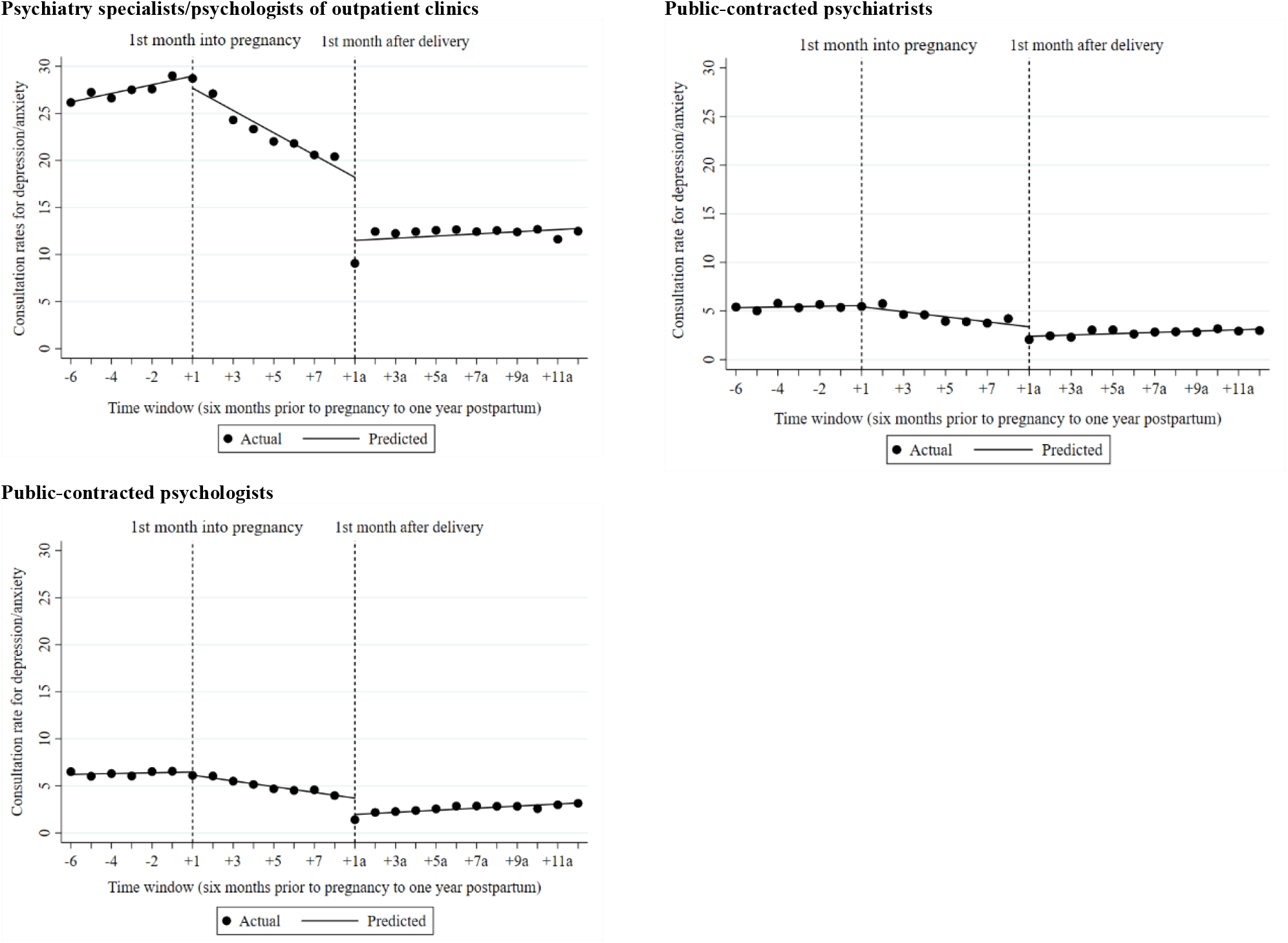
Consultation rate (number of consultations/100 pregnancies) for depression/anxiety with psychiatry specialists/psychologists of pregnant women having antidepressant fills for depression/anxiety prior to pregnancy: interrupted time-series analyses.

In the overall population, we observed an increase in consultation rate for depression/anxiety with psychiatric specialists/psychologists of outpatient clinics in the six months prior to pregnancy (slopes: 0.46 (95% confidence interval (CI): 0.23; 0.69)), notably in late discontinuers and interrupters while the consultations rates with public-contracted psychologists/psychiatrists remained stable (**Figure S3, S4, S5**).

Pregnancy start was associated with gradual decreases in the consultation rates for depression/anxiety (slopes during pregnancy: -1.18 (95%CI: -1.50; -0.87) for specialists of outpatient clinics, -0.26 (95%CI: -0.41; -0.11) for public-contracted psychiatrists, -0.31 (95%CI: -0.36; -0.25) for public-contracted psychologists) (see **Figure 1** and **Table 2**). The consultation rate during pregnancy of interrupters, however, remained stable (**Figure S3, S4, S5**).

**Table 2.** Consultation rate (number of consultations/100 pregnancies) for depression/anxiety with psychiatry specialists/psychologists of pregnant women having antidepressant fills for depression/anxiety prior to pregnancy: interrupted time-series analyses.

|  |  | Baseline<br>Coef (95% CI) | Slope prior<br>pregnancy<br>Coef (95% CI) | Change at first<br>month into<br>pregnancy<br>Coef (95% CI) | Slope during<br>pregnancy<br>Coef (95% CI) | Change at first<br>month after<br>delivery<br>Coef (95% CI) | Slope in the<br>postpartum year<br>Coef (95% CI) |
| --- | --- | --- | --- | --- | --- | --- | --- |
| **Psychiatric specialists/psychologists of outpatient clinics** |  |  |  |  |  |  |  |
|  | Overall | **26.19 (25.54; 26.84)** | **0.46 (0.23; 0.69)** | -1.29 (-2.98; 0.41) | **-1.18 (-1.50; -0.87)** | **-6.69 (-9.04; -4.35)** | 0.11 (-0.13; 0.36) |
|  | Late discontinuers† | **25.45 (24.49; 26.41)** | **0.98 (0.75; 1.21)** | -1.35 (-2.53; 0.17) | **-1.44 (-1.67; -1.20)** | **-9.67 (-11.59; -7.75)** | 0.01 (-0.19; 0.21) |
|  | Early discontinuers†† | **25.57 (23.20; 27.94)** | -0.13 (-0.76; 0.50) | -2.42 (-5.52; 0.69) | **-1.50 (-2.06; -0.93)** | -2.33 (-5.15; 0.50) | -0.002 (-0.21; 0.21) |
|  | Continuers††† | **29.77 (27.71; 31.82)** | 0.04 (-0.67; 0.76) | 1.15 (-2.11; 4.40) | **-1.03 (-1.28; -0.78)** | **-7.62 (-10.53; -4.72)** | 0.14 (-0.22; 0.50) |
|  | Interrupters†††† | **24.22 (22.36; 26.08)** | **1.06 (0.49; 1.62)** | -2.17 (-5.51; 1.18) | -0.20 (-0.68; 0.28) | **-7.56 (-11.45; -3.67)** | **0.54 (0.01; 1.06)** |
| **Public-contracted psychologists** |  |  |  |  |  |  |  |
|  | Overall | **6.23 (5.80; 6.65)** | 0.04 (-0.08; 0.16) | -0.33 (-0.77; 0.10) | **-0.31 (-0.36; -0.25)** | **-1.73 (-2.24; -1.22)** | **0.11 (0.05; 0.17)** |
|  | Late discontinuers | **4.92 (4.37; 5.48)** | 0.02 (-1.12; 0.17) | -0.47 (-1.08; 0.14) | **-0.17 (-0.30; -0.05)** | **-1.88 (-2.64; -1.12)** | 0.04 (-0.02; 0.11) |
|  | Early discontinuers | **5.56 (5.25; 5.86)** | **0.28 (0.17; 0.39)** | -0.70 (-1.47; 0.07) | **-0.42 (-0.47; -0.37)** | **-1.81 (-2.43; -1.19)** | **0.10 (0.02; 0.18)** |
|  | Continuers | **9.23 (5.91; 12.54)** | -0.28 (-1.16; 0.60) | 0.50 (-1.67; 2.68) | **-0.45 (-0.51; -0.39)** | **-2.24 (-2.73; -1.74)** | **0.22 (0.15; 0.29)** |
|  | Interrupters | **6.41 (5.91; 6.91)** | 0.06 (-0.16; 0.28) | -0.43 (-1.88; 1.01) | -0.17 (-0.38; 0.04) | -0.56 (-1.90; 0.79) | 0.13 (-0.01; 0.27) |
| **Public-contracted psychiatrists** |  |  |  |  |  |  |  |
|  | Overall | **5.35 (4.94; 5.74)** | 0.04 (-0.07; 0.14) | -0.12 (-0.78; 0.53) | **-0.26 (-0.31; -0.11)** | **-0.98 (-1.83; -0.12)** | **0.07 (0.02; 0.11)** |
|  | Late discontinuers | **4.72 (4.37; 5.08)** | 0.07 (-0.04; 0.20) | 0.15 (-0.80; 1.09) | **-0.36 (-0.54; -0.18)** | **-1.10 (-1.93; -0.27)** | **0.05 (0.03; 0.07)** |
|  | Early discontinuers | **5.04 (4.51; 5.48)** | -0.02 (-0.14; 0.11) | **-1.24 (-1.87; -0.62)** | **-0.22 (-0.36; -0.08)** | -0.67 (-1.58; 0.24) | 0.03 (-0.02; 0.07) |
|  | Continuers | **6.70 (6.16; 7.23)** | -0.13 (-0.25; -0.01) | **1.56 (0.71; 2.40)** | **-0.31 (-0.52; -0.10)** | -1.22 (-2.46; 0.02) | 0.01 (-0.05; 0.07) |
|  | Interrupters | **5.50 (4.42; 6.58)** | **0.29 (0.01; 0.57)** | -0.75 (-1.74; 0.25) | -0.04 (-0.18; 0.09) | -1.00 (-2.25; 0.26) | **0.25 (0.09; 0.41)** |

Abbreviations: coef – coefficient, CI – confidence interval. Significant estimates are indicated in **bold**. Slope prior to pregnancy reflects the change per month in the consultation rate in the six months prior to pregnancy (e.g., slope = 9.1 indicating a gradual increase of 9.1 consultations per 100 pregnancies each month prior to pregnancy). Change at the first month into pregnancy refers to the immediate change associated with the start of pregnancy. Slope during pregnancy indicates the trend in the consultation rate in the eight months into pregnancy. The unit of these measures was the number of consultations per 100 pregnancies
†late discontinuers referred to those discontinued antidepressant treatment around third trimester ††early discontinuers referred to those discontinued antidepressant treatment around the start of pregnancy
†††continuers referred to those continued antidepressant treatment throughout pregnancy and postpartum year and ††††interrupters referred to those discontinued antidepressant treatment in late pregnancy and resumed in postpartum year

On the other hand, the delivery was associated with immediate drop but gradual increase in consultation rate with public-contracted psychologists/psychiatrists (slopes: 0.11 (95%CI: 0.05; 0.17) and 0.07 (95%CI: 0.02; 0.11)) while the consultation rate with specialists of outpatient clinics dropped following delivery then remained stable afterwards. The increase in the consultation rates for depression/anxiety of interrupters with specialists of outpatient clinics and public-contracted psychiatrists were more pronounced than other trajectories (see **Table 2 and Figure S3, S4, S5**).

### Sensitivity analyses

Similar to psychiatric specialists/psychologists, we also observed a decrease in the consultation rate for depression/anxiety with GPs when pregnant women entered their course of pregnancy, and likewise a rate increase in the postpartum year in the overall population (**Figure S6**) and across all antidepressant fill trajectories (data not shown).

The analyses limited to one pregnancy per woman and with extended inclusion criteria yielded consistent findings (data not shown).

The analyses on consultation rates with GPs and physicians of outpatient clinics for any conditions showed different patterns compared to depression/anxiety: increase during pregnancy and decrease in the postpartum year (**Figure S7**).

## DISCUSSION

### Summary of findings

This nationwide study describes mental health care utilization of a cohort of pregnant women with depression/anxiety prior to pregnancy comprising nearly 9,000 pregnancies. First, we observed a reduced mental health care utilization when pregnant women entered the course of pregnancy. Notably, the declines were observed not only among women who continued their antidepressant treatment throughout their pregnancy but also among those who discontinued their treatment in pregnancy. Second, we found increases in mental health care utilization in the postpartum year, notably with public-contracted psychiatrists/psychologists. The increase was more pronounced in interrupters who resumed the treatment after discontinuation. Third, consultations for depression/anxiety with public-contracted psychiatric specialists accounted for one third of total care with psychiatric specialists.

### Interpretations

Little attention has been paid to the use of alternative non-pharmacological care to substitute antidepressant treatment among those who discontinued their pharmacologic treatment. The majority of our population (nearly 80%) discontinued their antidepressant treatment in early or late pregnancy. Because most antidepressant prescriptions were issued by GPs, consultations for depression/anxiety with GPs which were likely for prescription renewal also decreased during pregnancy. Consultations for depression/anxiety with psychiatric specialists and psychologists which are mostly for psychotherapy and psychiatric follow-up, followed the same patterns. The decreases were observed across all antidepressant fill trajectories suggesting that antidepressant treatment was less likely to be replaced by psychotherapy, notably among discontinuers. Approximately one third of those who discontinued antidepressant use during the third trimester progressively resumed antidepressant treatment after delivery, suggesting potential re-occurrence or relapse of depression/anxiety after antidepressant discontinuation. Indeed, the consultation rate for depression/anxiety with specialists of outpatient clinics and public-contracted psychiatrists increased after delivery among interrupters. One previous study performed in French setting found that about 22% of women who discontinued antidepressant treatment after conception resumed their treatment later in pregnancy. This interruption pattern suggests a lack of compliance in pregnancy due to fear of teratogenic drug effects on the unborn child and/or a potential relapse due to discontinuation.^13^ The interrupters in our population, however, started to resume their treatment after delivery potentially because reinitiating antidepressants in late pregnancy is not recommended.^4^ Unlike non-pregnant women, pregnant women might also consult with their obstetricians and gynaecologists for depression/anxiety, which potentially explains the decrease in the consultations with GPs and psychiatric specialists for these conditions. However, we identified very few consultations with obstetricians and gynaecologists having a reimbursement code for depression/anxiety.

In Norway, all pregnant and postpartum women are offered public follow-up by a midwife at the maternal and child health centres. These centres offer both individual and group-based support for families, and may have access to psychologists (not mandatory) for early prevention and treatment for perinatal mental disorders.^27^ Based on the above and our own results, we cannot exclude the possibility that the reduced mental health care utilization at specialist care level may be counterbalanced by a greater uptake at the maternal and child health centres, which are more easily accessible and have no waiting time. Yet, psychologist and perinatal mental health specialists may not always be available at the centres. To safeguard maternal mental health, as well as child and family well-being, it is therefore crucial to establish a systematic cooperation among primary-level and between-level health professionals, so that women with depression/anxiety with different antidepressant treatment preferences, are adequately followed-up and treated.

Our sensitivity analyses also showed that decreased consultation rates during pregnancy and increased consultation rates after delivery were specific for depression/anxiety. Future studies should explore whether or not the same patterns were observed in other countries and address whether or not the decline in mental health care utilization reflects undertreatment among pregnant women presented with depression and/or anxiety.

### Strengths and limitations

To our knowledge, the present study is the first study to investigate the mental health care utilization regarding different antidepressant fill trajectories in pregnant women with depression/anxiety. We used data retrieved from various nationwide registries that provide insights of a representative cohort with exhaustive recording on both medication fills and outpatient health care utilization (including both primary and secondary care). The availability of diagnostic codes in ICD-10 and ICPC-2 allowed us to get a specific targeted population having depression and/or anxiety.. The use of trajectory modelling allowed us to overcome the simple categorization of continuers and discontinuers. Several sensitivity analyses were performed and confirmed the robustness of our findings.

We noted some limitations in this study. First, we could not investigate whether or not antidepressants were discontinued by the patient or the prescribing physician. Moreover, it was not possible to assess whether the patient was undertreated or not. Second, information on indications and prescribers were not fully provided in the NorPD and the exact proportion of antidepressants filled for depression and/or anxiety is unknown. However, our study population included only pregnancies within women having received a clinical diagnosis for depression and/or anxiety and the majority of antidepressant prescriptions have depression and/or anxiety as an indication.^8^ Third, we used consultation rate with psychiatric specialists as a proxy for non-pharmacological care which might have resulted in overestimation of the non-pharmacological care in pregnant women. However, specialists issued only 10% of antidepressant prescriptions while the number of consultations for depression/anxiety are similar to those of GPs. However, psychiatrist specialists may often counsel pregnant women in relation to antidepressant treatment even though they do not directly prescribe these medications. In addition, we assessed public-contracted psychologists as a separate group because they can only offer psychotherapy but not medication and this group had similar patterns with other types of psychiatric specialists. Fourth, we were unable to differentiate psychologists and psychiatrists of outpatient clinics in our data. Because the waiting time and costs related to consultations with specialists of outpatient clinics and public-contracted specialists are similar, consultation patterns with public-contracted specialists can be used as proxy for those of specialists of outpatient clinics. Fifth, the actual use and pattern of use of antidepressants after being filled is not recorded in our databases. Previous studies show high agreement between antidepressant fills in NorPD and self-reported antidepressant use in the Norwegian Mother and Child Cohort Study for the window of exposure from gestation week 21 to delivery.^28^ Sixth, antidepressant use in the hospital was not covered within this study. Our preliminary analyses showed that a very small proportion of the study population was hospitalized for a diagnosis of depression and/or anxiety. Sixth, in Norway, the waiting time to have a consultation with specialists in public service (i.e., outpatient clinics) or with a contract specialists is considerably long (maximum allowed waiting time can be up to 14 weeks after referral of GPs), and patients may need to seek for care with private psychiatric specialists with a potential shorter waiting time.^27^ However, the activities of private psychiatric specialists were not covered in KUHR and NPR. Last but not least, the ITSA is robust to measure the potential impact of pregnancy on the patterns of outcome measures but the assumption of fixed interruption points did not provide us with the best fitted models for the actual data.

## Conclusions

Pregnancy was associated with reduced mental health care utilization regardless of whether antidepressant treatment was maintained during pregnancy or not. An increase in mental health care utilization in the postpartum year was observed among those pregnancies within women who discontinued their antidepressant treatment during pregnancy then resumed after delivery.

## Data Availability

The data in this project were delivered by the registry holders to the researchers as pseudonymized data files. Data are available upon request to the registry holders, provided legal and ethical approvals.

## AUTHORS’ CONTRIBUTIONS

Conception of study: NT, AL

Data access and approvals: HN

Design of study: NT, HN, GB, MEG, AL

Data management: NT

Data analysis: NT

Drafting of the manuscript: NT, AL

Revising of the manuscript: HN, GB, MEG

Final approval of the version to be published: NT, HN, GB, MEG, AL

## ACKNOWLEDGEMENT

Data was stored at the TSD (Tjeneste for Sensitive Data) facilities, owned by the University of Oslo, operated and developed by the TSD service group at the University of Oslo, IT-Department (USIT).

We are grateful for constructive comments from Dr Kristin Palmsten on the earlier version of the manuscript.

Nhung Trinh and Angela Lupattelli are supported by the Norwegian Research Council (grant no. 288696). Hedvig Nordeng is supported by a European Research Council Starting Grant DrugsInPregnancy (grant number 639377).

No financial relationships with any organizations that might have an interest in the submitted work in the previous three years, no other relationships or activities that could appear to have influenced the submitted work. The funders had no role in the design and conduct of the study; the analysis and interpretation of the data; preparation; the review or approval of the manuscript.

## ETHICS

The study was approved by the Regional Committee for Research Ethics in South Eastern Norway (approval number 2018/140/REK Sør Øst) and by the Data Protection Officer at the University of Oslo (approval number 58033). Data were handled in accordance with the General Data Protection Regulation.

## TRANSPARENCY DECLARATIONS

The authors have no conflict of interest to declare. We affirm that this manuscript is an honest, accurate, and transparent account of the study being reported; that no important aspects of the study have been omitted; and that any discrepancies from the study as planned (and, if relevant, registered) have been explained. The findings and conclusions in this report are those of the authors and do not necessarily represent the official position of any organization or company.

