## Appendices for "Antidepressant and mental health care utilization in pregnant women with depression and/or anxiety: an interrupted time-series analysis"

Figure S1. Flowchart of study population

Figure S2. Trajectories of antidepressant prescription fill in pregnant women having antidepressant fills for depression/anxiety prior to pregnancy: Solid lines representing the main trajectories and the dotted lines representing the 95% confidence interval of the corresponding trajectories.

Figure S3. Consultation rate (number of consultations/100 pregnancies) for depression/anxiety with psychiatry specialists/psychologists of outpatient clinics of pregnant women having antidepressant fills for depression/anxiety prior to pregnancy: interrupted time-series analyses

Figure S4. Consultation rate (number of consultations/100 pregnancies) for depression/anxiety with public-contracted psychiatrists of pregnant women having antidepressant fills for depression/anxiety prior to pregnancy: interrupted time-series analyses

Figure S5. Consultation rate (number of consultations/100 pregnancies) for depression/anxiety with public-contracted psychologists of pregnant women having antidepressant fills for depression/anxiety prior to pregnancy: interrupted time-series analyses

Figure S6. Consultation rate for depression/anxiety with general practitioners of pregnant women having antidepressant fills for depression/anxiety prior to pregnancy: interrupted time-series analyses

Figure S7. Consultation rate for all conditions with general practitioners and specialists of outpatient clinics of pregnant women having depression/anxiety prior to pregnancy: interrupted time-series analyses

**Figure S1. Flowchart of study population**

MBRN pregnancies

2009-2018

n=592,189

Missing maternal ID: n=0

Missing or invalid gestational length (>45 weeks): n=2,523

Gestational length less than 32 weeks: n=9,727

MBRN pregnancies linked to NorPD, KUHR, NPR

2009 - 2018

n=579,939

No antidepressant filling or no encounter for depression and/or anxiety in the six months before pregnancy: n=571,149

MBRN pregnancies linked to NorPD, KUHR, NPR

2009 - 2018

n=8,520

**Full study cohort**

MBRN pregnancies linked to NorPD, KUHR, NPR

2009 - 2018

n=8,460
Among 8,092 women

Gap between pregnancies less than one year: n=60

MBRN, Medical Birth Registry of Norway; NorPD, Norwegian Prescription Database; KUHR, Norway Control and Payment of Health Reimbursement (KUHR); NPR, Norwegian Patient Registry

**Figure S2. Trajectories of antidepressant prescription fill in pregnant women having antidepressant fills for depression/anxiety prior to pregnancy: Solid lines representing the main trajectories and the dotted lines representing the 95% confidence interval of the corresponding trajectories.**

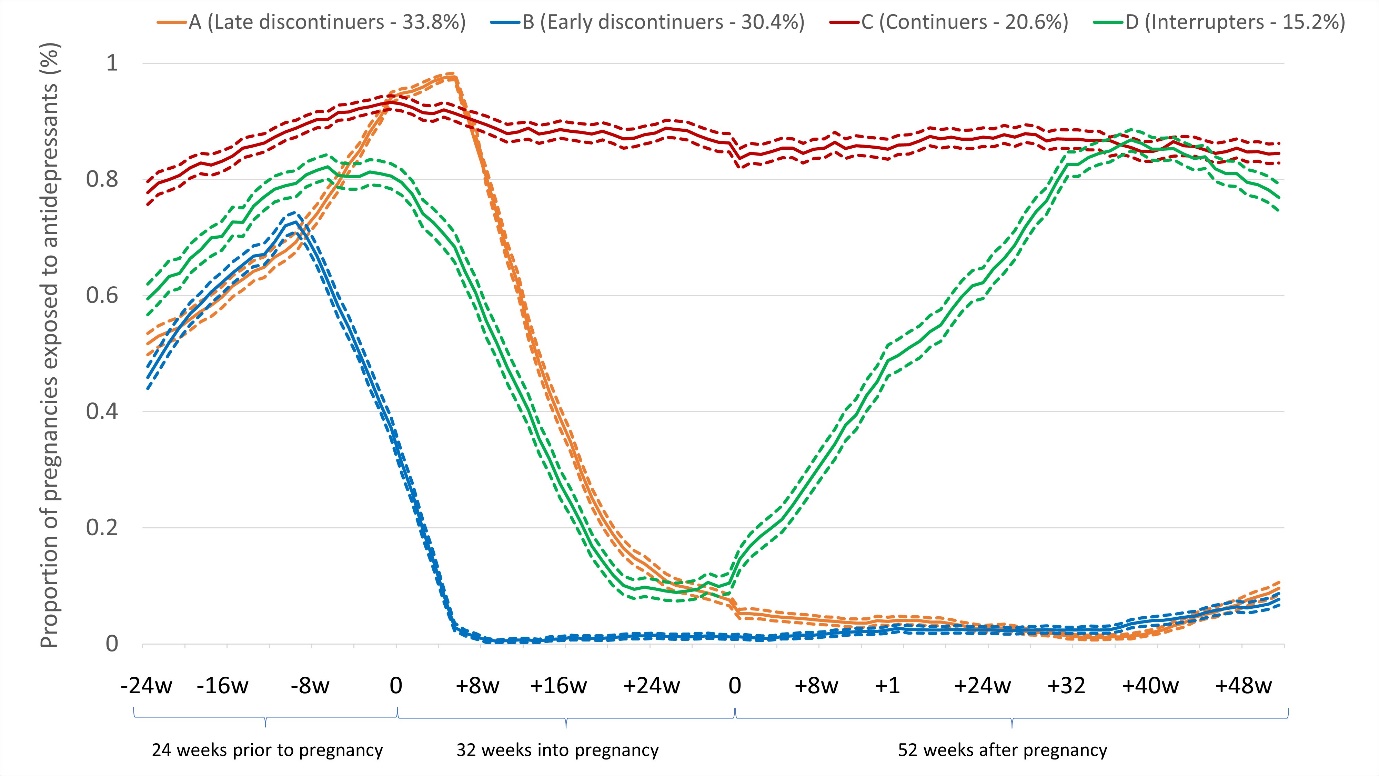

**Figure S3. Consultation rate (number of consultations/100 pregnancies) for depression/anxiety with psychiatry specialists/psychologists of outpatient clinics of pregnant women having antidepressant fills for depression/anxiety prior to pregnancy: interrupted time-series analyses**

| **Late discontinuers**  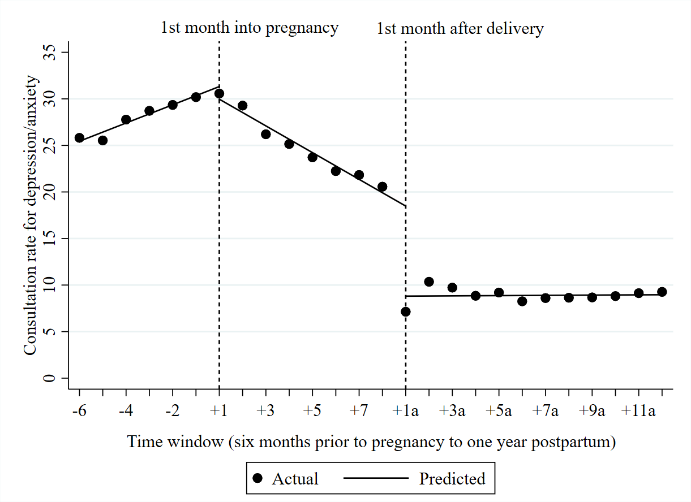 | **Early discontinuers**  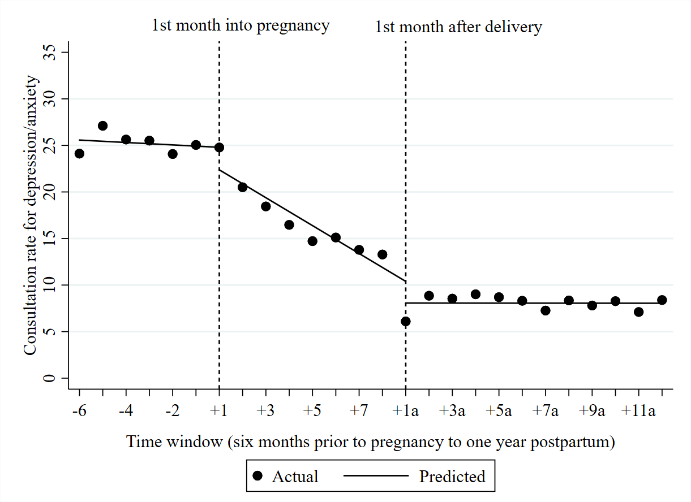 |
| --- | --- |
| **Continuers**  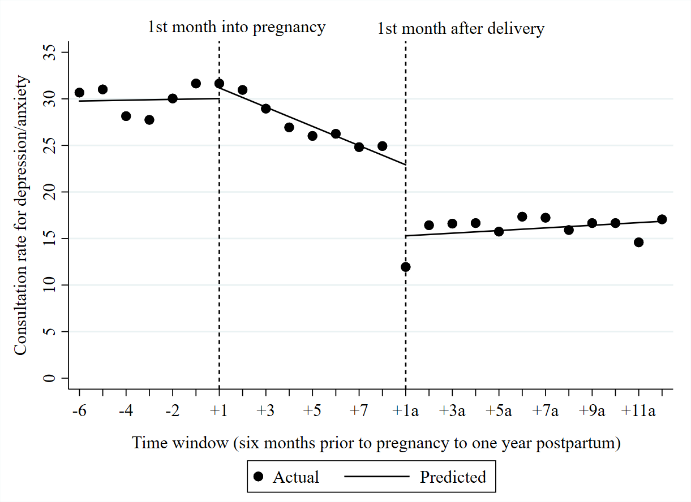 | **Interrupters**  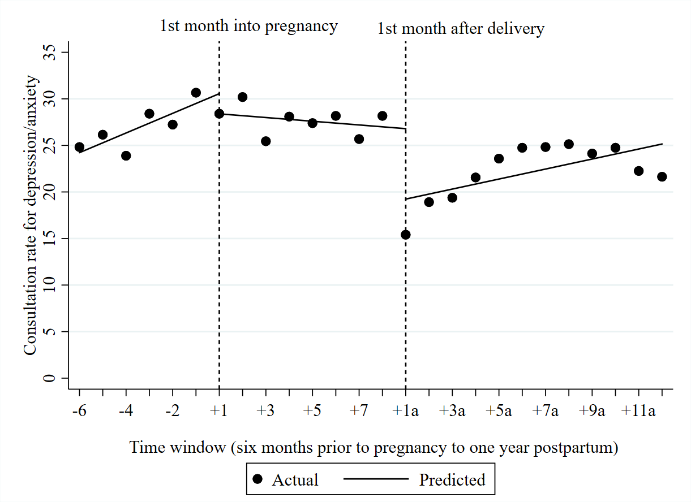 |

**Figure S4. Consultation rate (number of consultations/100 pregnancies) for depression/anxiety with public-contracted psychiatrists of pregnant women having antidepressant fills for depression/anxiety prior to pregnancy: interrupted time-series analyses**

| **Late discontinuers**  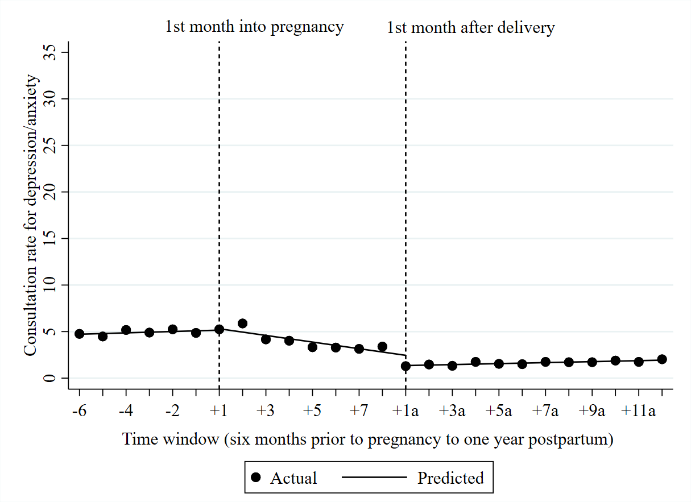 | **Early discontinuers**  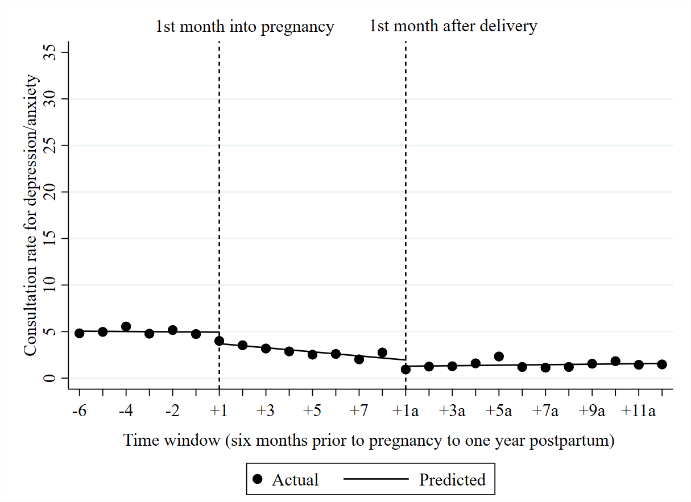 |
| --- | --- |
| **Continuers**  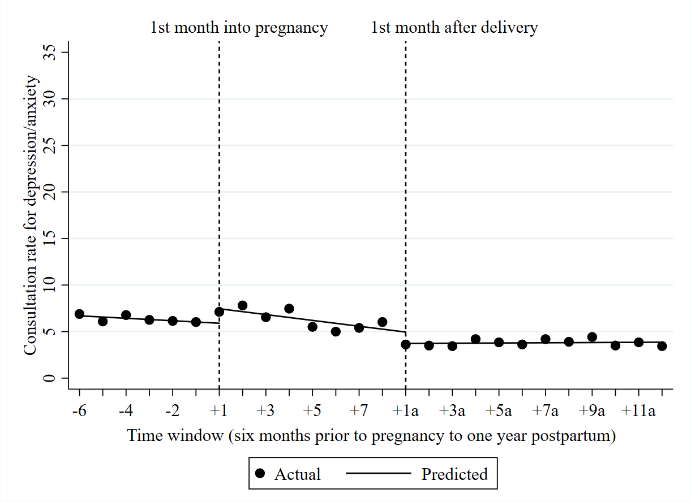 | **Interrupters**  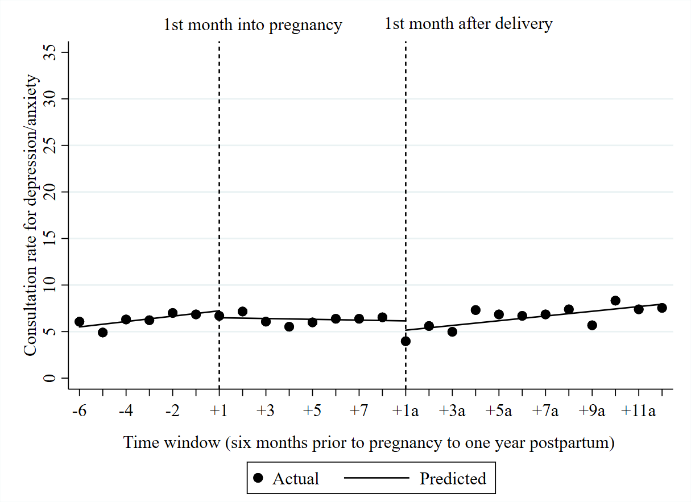 |

**Figure S5. Consultation rate (number of consultations/100 pregnancies) for depression/anxiety with public-contracted psychologists of pregnant women having antidepressant fills for depression/anxiety prior to pregnancy: interrupted time-series analyses**

| **Late discontinuers**  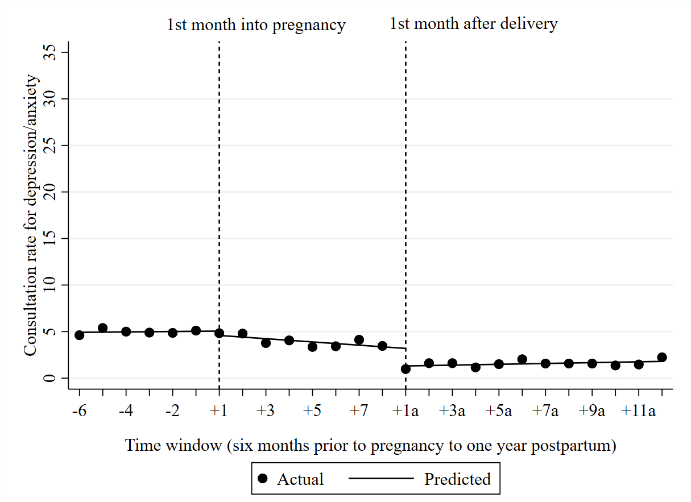 | **Early discontinuers**  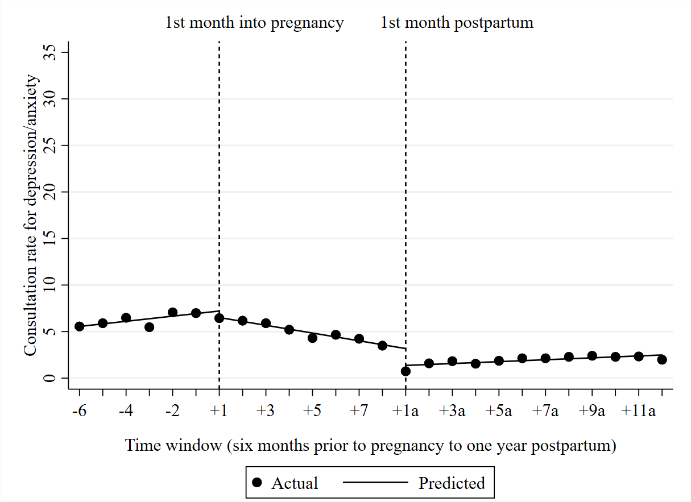 |
| --- | --- |
| **Continuers**  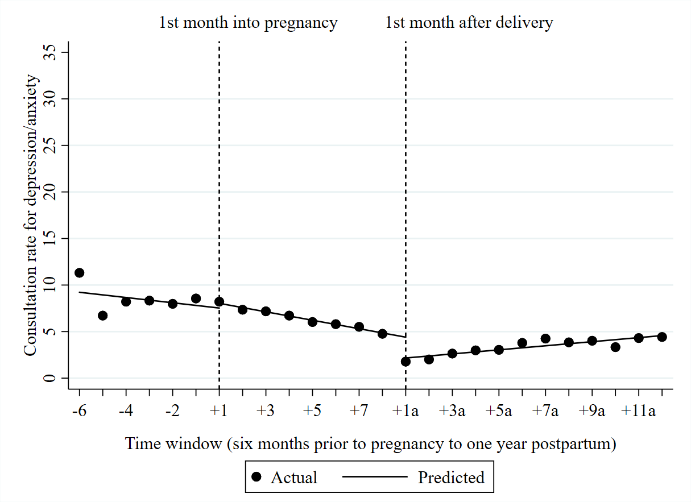 | **Interrupters**  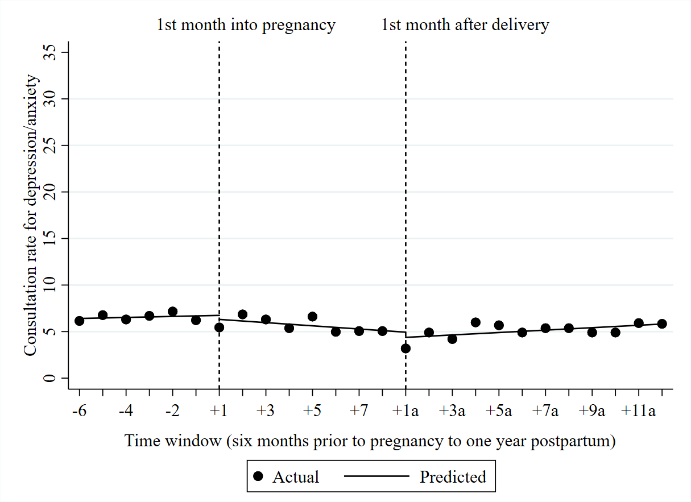 |

**Figure S6. Consultation rate (number of consultations/100 pregnancies) for depression/anxiety with general practitioners of pregnant women having antidepressant fills for depression/anxiety prior to pregnancy: interrupted time-series analyses**

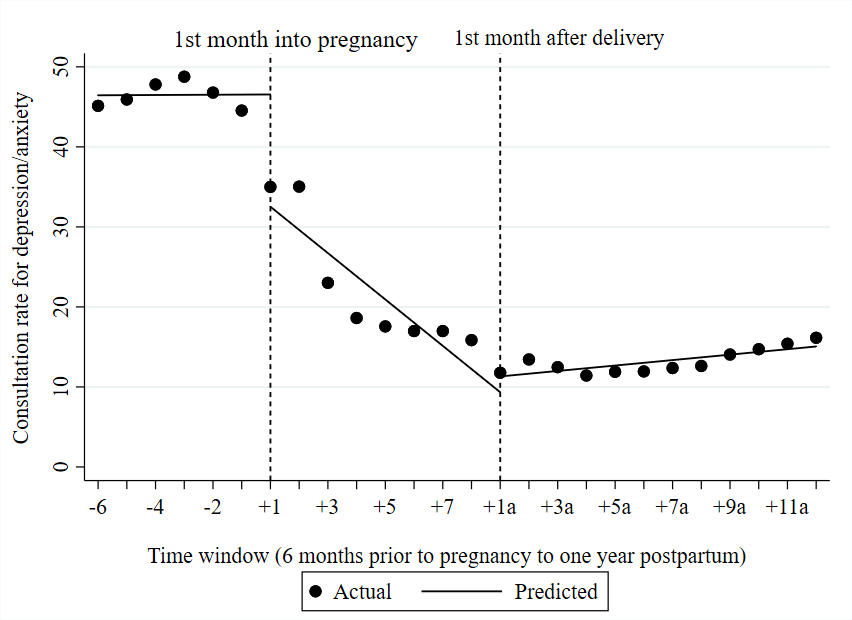

**Figure S7. Consultation rate (number of consultations/100 pregnancies) for all conditions with general practitioners and specialists of outpatient clinics of pregnant women having depression/anxiety prior to pregnancy: interrupted time-series analyses**

**General practitioners**

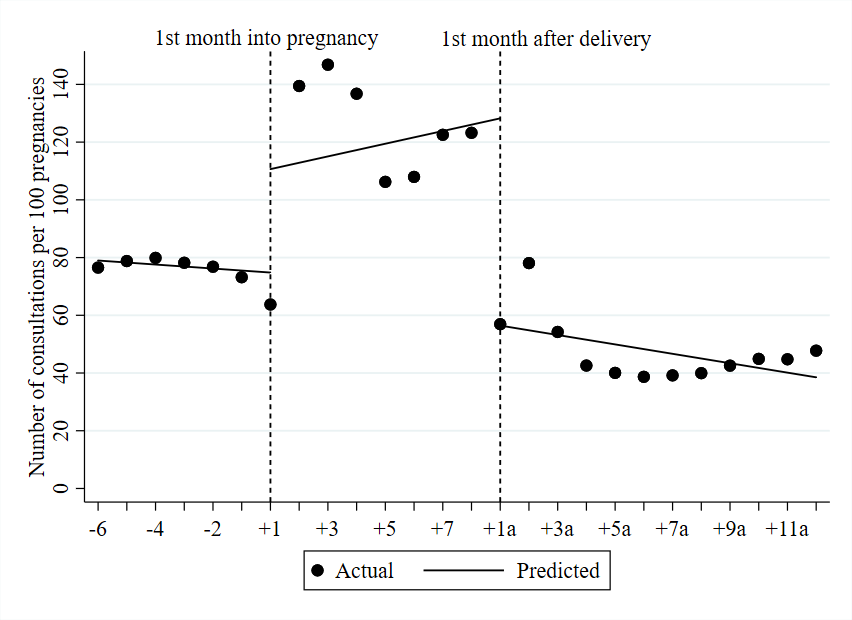

**Specialists of outpatient clinics**

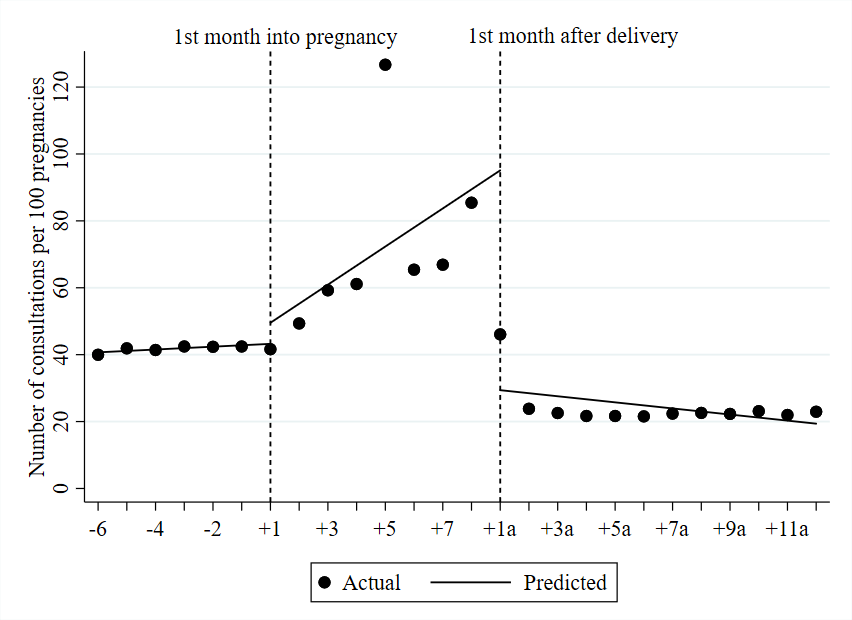
